# Tracking Closely Related Enteric Bacteria at High Resolution in Fecal Samples of Premature Infants Using a Novel rRNA Amplicon

**DOI:** 10.1101/2020.09.26.20201608

**Authors:** J. Graf, N. Ledala, M. J. Caimano, E. Jackson, D. Gratalo, D. Fasulo, M. D. Driscoll, S. Coleman, A. Matson

## Abstract

Identifying and tracking microbial strains as microbiomes evolve is a major challenge in the field of microbiome research. Longitudinal microbiome samples of co-admitted twins from two different neonatal intensive care units (NICUs) were analyzed using a ∼2,500 base amplicon that spans the 16S and 23S rRNA genes, and mapped to a new 16S-23S rRNA database. Amplicon Sequence Variants inferred using DADA2 provided sufficient resolution for differentiation of rRNA variants from closely related, but not previously sequenced *Klebsiella, E. coli*, and *Enterobacter*, among the first bacteria colonizing the gut of these infants after admission to the NICU. Distinct ASV groups (fingerprints) were followed between co-admitted twins over time, demonstrating the potential to track the source and spread of both commensals and pathogens.The high-resolution taxonomy obtained from long amplicon sequencing enable tracking of strains temporally and spatially as microbiomes are established in infants in the hospital environment.

## Introduction

The gut microbiome helps establish and maintain critical systems required for lifelong health, including the establishment and maintenance of the gut mucosal barrier, endocrine system, nutritional metabolism, and pathogen defense (*1*). Understanding factors that influence the establishment of the gut microbiome requires a detailed understanding of the diverse array of microbial strains involved during development (*2*). Most publications to date rely on sequencing a short region of the 16S rRNA gene that typically allows for identification to the family or genus level but only in rare cases to the species or strain level (*3*). The importance of strain level identification is exemplified by *Escherichia coli*, which includes benign strains, as well as pathogenic strains, such as serotypes O104:H4 and O113:H21, which carry the Shiga toxin gene responsible for hemorrhagic diarrhea (*4*). Since strains belonging to the same species can range from benign to pathogenic, strain-level identification is an important barrier that needs to be overcome in order to decipher links between the microbiome and human health.

DNA sequencing techniques are useful for both high and low resolution taxonomic identification of bacteria in microbiome samples. The ability to differentiate closely related bacteria using DNA sequencing methods depends on the read length and quality of DNA sequence obtained, as well as the taxonomic curation and depth of content of the reference database used for mapping. In addition, the accuracy of the software algorithms used to compare the sequence data with reference databases or infer amplicon sequence variants may vary (*5*). In general, increasing amplicon read length is useful in obtaining higher resolution taxonomy for bacteria in a microbiome (*6*). Since the genome of a single bacterium contains multiple 16S-23S rRNA genes that can vary in sequence, the combination and relative abundance of 16S-23S variants can be useful in determining bacterial taxonomy at the species level and lower. For example, the *E. coli UMN026* genome contains seven copies of the 16S and 23S genes, separated by a short internally transcribed spacer (ITS) sequence between that can vary in length (Fig. 1). The seven copies produce seven distinct amplicons for a single genome, which vary in length and sequence. Generating multiple amplicons per genome presents the opportunity to use the combination as a ‘fingerprint’ profile to identify a given strain, even if closely related strains share one or more 16S-23S variants. In this study, a high-resolution ∼2,500 base pair 16S-ITS-23S amplicon (StrainID) was used to explore whether 16S-23S amplicons, and combinations thereof, from *Klebsiella* spp., *E. coli*, and *Enterobacter* spp. present in premature infant fecal samples could be used to identify and track individual strains during microbiome development.Amplicon Sequence Variants (ASVs) were inferred using DADA2 (*7*), and used to confirm the presence of previously unsequenced strains, and to determine whether these novel organisms were present in different longitudinal samples and individuals. The results demonstrate that 16S-23S StrainID amplicon fecal microbiome sequencing generates multiple distinct ASVs for individual bacterial strains, and that the combination and relative abundance of ASVs can be used as a fingerprint to track previously unsequenced species both longitudinally across time and across individuals in the NICU.

**Fig 1.**
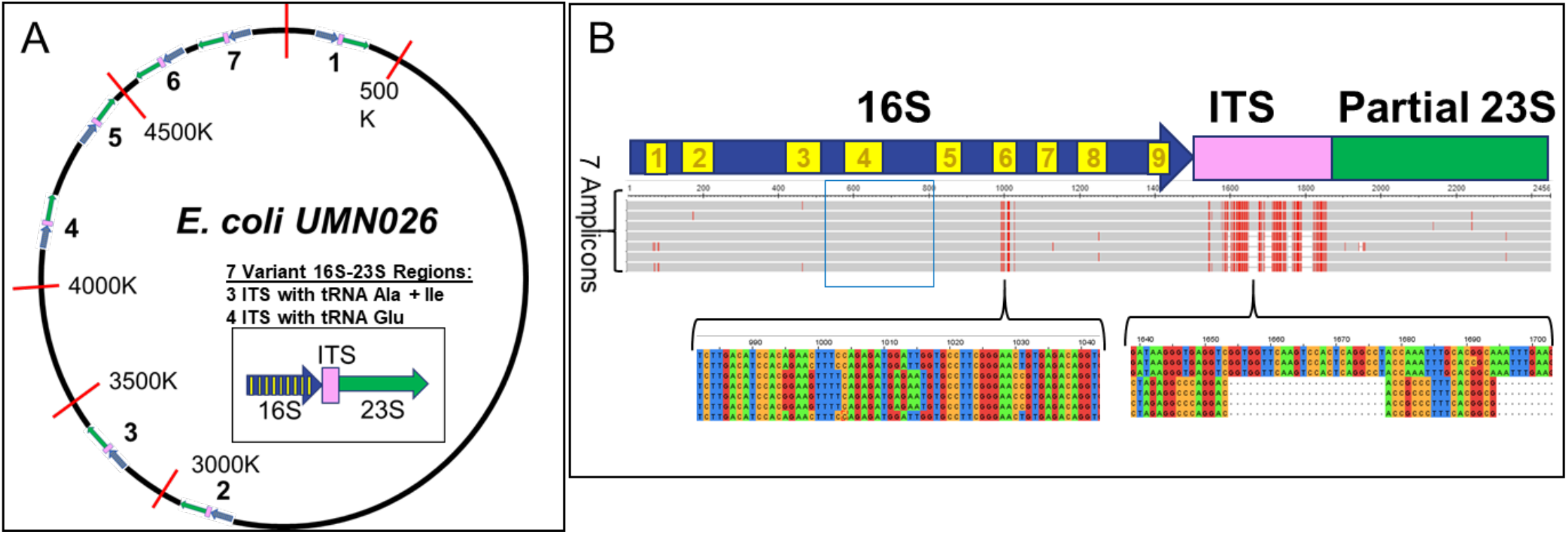
A single *E. coli* genome can generate multiple StrainID amplicons. **(A)** A circle representing the *E. coli UMN026* genome, with approximate placement of the seven 16S-23S rRNA gene pairs is shown. The Internally Transcribed Spacer between the genes is represented as a pink box, the 16S rRNA gene is a blue arrow, and the 23S rRNA gene is represented by a green arrow. **(B)** Multiple alignment of the 7 amplicon regions. The sequences from *E. coli UMN026* NC_011751 were aligned using Jalview and visualized in Multiple Sequence Alignment Viewer 1.14.0 (https://www.ncbi.nlm.nih.gov/projects/msaviewer/). The blue arrow with the numbered yellow boxes represents the approximate position of the 16S rRNA gene in the aligned sequences, with the numbered yellow boxes indicating the approximate locations of the nine 16S variable regions. The red hash marks in the gray aligned sequences indicate variant bases. The white gaps indicate deletions. Three of the seven 16S-23S amplicons contain two tRNA genes, tRNA Ala and tRNA Ile, in the ITS, while the four shorter ITS contain a single tRNA Glu. The base level alignments at the bottom detail some of the key variant sites in the V6 region and in the ITS region indicated by brackets. The blue box surrounding the 16S V4 highlights an invariant sequence across all 7 amplicons that includes bases 513-806 in the 16S V4 region that is a common 16S target amplicon region.

## Results

### StrainID amplicon enables species and strain level detection of pathogens in fecal microbiomes from premature infants

Sequential fecal samples from two sets of premature twins, born with gestational ages of 30 and 29 weeks and cared for in two different NICUs, were collected and analyzed. Figure 2 demonstrates that infant twins A and B shared *Klebsiella* spp., *E. coli*, and *Enterobacter* spp., which comprised more than half of the reads recovered from every sample. *Enterobacter cloacae* strains, shown with red border, are a common pathogen in NICU environments (*8*), and were present at all time points. *Klebsiella pneumoniae* strains, shown with black border, appeared in Twin A at week 4, and Twin B at week 3, and remained a significant component of the overall read count through week 8 in both twins. *E. coli* strains, with light blue border, were present in Twin A by week 3 and Twin B by week 4, and also remained established through week 8.Microbial diversity increased over time, with the appearance of the common digestive-tract residents *Veillonella* spp. and *Bifidobacterium* spp. at later time points. The important gut symbiont *Bifidobacterium longum* (*9*) (pink box) is established early in Twin A, present in small quantity in week 3, remaining over 5% of reads through week 8. *Bifidobacterium longum* is established in Twin B only at the final week 8 time point. It is interesting to speculate on the source of *B. longum* in Twins A/B, because *B. longum* did not appear as a substantial portion of the microbiome in the Y/Z twin pair, as described below. For the first 4-5 weeks, both twin pairs received a diet consisting primarily of human milk, an important source of oligosaccharides that benefit the growth of *Bifidobacterium* spp.(*10*); at later weeks, the human milk diet was supplemented with formula as the infants neared discharge.

**Fig 2.**
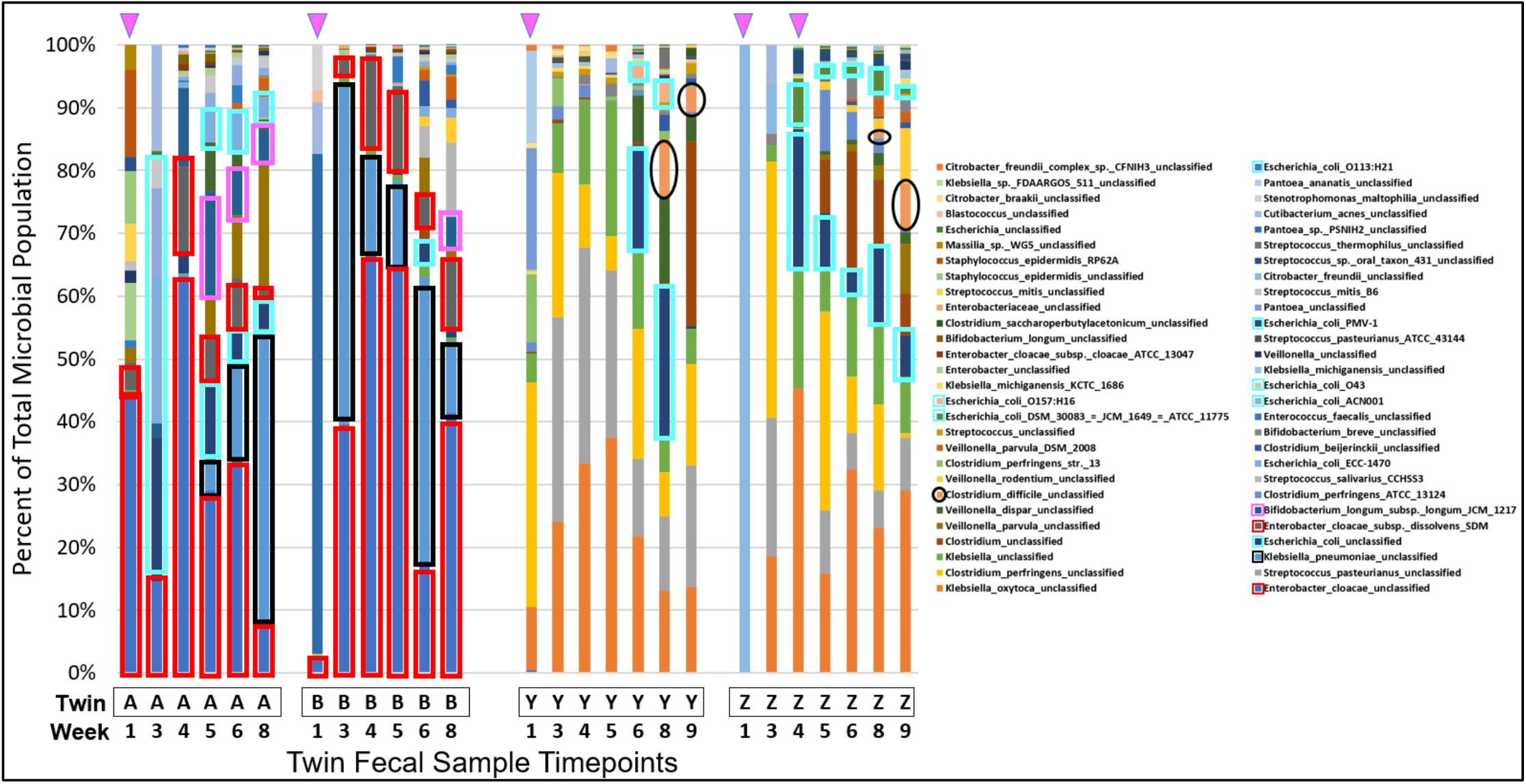
Twin Pairs A/B and Y/Z Share Similar Bacterial Profiles Within but Not Between Pairs. Fecal samples from two pairs of twins were profiled from week 1 to week 8 (A/B) or 9 (Y/Z) after birth. Twin pair A/B and Y/Z were born at 30 and 29 weeks, respectively. Twins A/B were admitted to a NICU in a different hospital than Twins Y/Z. Samples were obtained during the week indicated on the x-axis. Results are shown for each sample as a ‘100% Stacked Column’, with Y-axis box size indicating percent of the total microbial population. Pink triangles at the top of the Figure indicate antibiotic treatment prior to sample collection. Specific taxa were highlighted with the indicated colored border *Enterobacter cloacae* (red), *Klebsiella pneumoniae* (black) *E. coli* taxa (light blue), *Bifidobacterium longum* (pink), and *Clostridium difficile* (tan bars with black ovals)

The major bacterial species were shared within each pair of twins, but there were striking differences between the twin pairs. As with A/B, there is a high degree of similarity within the bacterial species present in the Y/Z twin pair. There were only 10 reads recovered from the week 1 sample for Twin Z, so that sample was excluded from further analysis. For all other samples in Twins Y and Z, *Klebsiella oxytoca* (orange bars) and *Clostridium perfringens* (yellow bars) were present at high levels, both of which have been associated with necrotizing enterocolitis in preterm infants (*11, 12*). Many of the samples also included high levels of *Streptococcus pasteurianus* (gray bars), a Group D *Streptococcus* sp. reported to be a cause of sepsis and meningitis in twin infants (*13*). Interestingly, absence of *S. pasteurianus* corresponded with empiric treatment with ampicillin and gentamicin for both Twins Y/Z at week 1, and following a second course of the same antibiotics for suspected sepsis in Twin Z between weeks 3 and 4; however, it reappeared in Twin Z by the following week and was present at all subsequent time points. A similar pattern was observed for *Clostridium perfingens*, which decreased below the level of detection in Twin Z at week 4 following antibiotic treatment, but reappeared by week 5 and was subsequently maintained. Interestingly, *Klebsiella* would normally be expected to be affected by treatment with gentamicin, but instead increased as a proportion of the microbiome. However, the week 4 time point also had a decreased number of reads, an indication that overall bacterial load may have been decreased, so it is possible that absolute numbers of *Klebsiella* decreased at a lower rate, leading to a greater proportion of the overall representation. *E. coli* appeared in Twin Z by week 4 concurrent with antibiotic treatment, but was not present in Twin Y until week 6, an indication that antibiotics may have facilitated more rapid colonization from low to undetectable levels by reducing microbial competition. *Clostridium difficile* (black circles) appeared at weeks 8-9 for both Twins Y/Z.

As premature infant samples harbor a low complexity microbiome, relatively few reads were needed for accurate representation of the most abundant organisms. Shannon diversity plots for Twin Y illustrate that fewer than 50 reads were required to converge on the relatively low number of taxa in each of the samples, even as the species diversity increased (Supplementary Fig, S1). Weeks 1-3 and time points with antibiotic treatment tended to correspond with recovery of fewer reads/sample. For example, there were fewer than 500 reads at weeks 1 and 3 for Twins A/B, week 1 for Twin Y, and week 1, 3 and 4 (antibiotics) for Twin Z. However, for most samples with few reads, the organisms represented were consistent with those in later time points. The initial samples which largely represent meconium stool that develops during the fetal period, or the early provision of antibiotics depleting populations of colonizing microbes, could be the cause of limited bacterial load and low recovery of DNA in specific samples. It is noteworthy that pathogens found commonly within hospital environment were among the first bacteria to colonize the preterm infant gut, and these were maintained at later time points, suggesting a role of the built environment in seeding the microbiome of premature infants.

These initial mapping data demonstrate that high resolution taxonomic assignments enable tracking of specific gut bacteria including important pathogens during the colonization of the premature infant gut within the NICU environment at species and even strain-level resolution. As bacterial communities are established and diversify within individuals, the ability to track the ebb and flow of specific strains provides an important tool for correlating bacterial colonization with health outcomes. This approach may also provide a means for assessing methods for accelerating the uptake common gut bacteria and replacement of early colonizers that may include NICU pathogens in the infant microbiome. However, the mapping data also revealed that there were a high number of reads that were labelled ‘unclassified’ at the species or strain level. Since species or strains can differ in sequence by only a few nucleotides, and rare sequencing errors can be present in even the most accurate sequencing data, error corrected reads are needed to determine whether the reads are ‘unclassified’ because they are from novel strains, or if the mapping was poor due to sequencing error. To facilitate high-resolution taxonomic assignments of closely related ‘unclassified’ novel bacteria, the sequence data were analyzed using DADA2 to infer Amplicon Sequence Variants (*7*). The ASVs represent the exact sequences of the 16S-23S amplicons from bacteria in the samples, enabling high-resolution comparison at the single base level to known genomes and other ASVs.

### DADA2 was used to infer Amplicon Sequence Variants (ASVs) from Klebsiella reads present in the samples

Sequencing-related methods to track *E. cloacae* infections have been shown to be useful in tracking strain transmission (*14*), but given the number of ‘unclassified’ sequences in these samples, and the possibility that strains can differ by only a few bases, it was clear that comparing closely related *Enterobacter* spp., *E. coli*, and *Klebsiella* spp. by mapping amplicon reads to the 16S-23S (Athena) database alone would be insufficient for strain tracking. To move beyond the limitations of mapping individual reads to reference sequences, we used DADA2 to generate ASVs from the StrainID sequencing data. ASVs inferred using DADA2 are StrainID amplicons that are correct at the single base level. The ASVs sequences can be compared not only to the reference genomes, but also against each other, to determine if the sample contains novel ASVs that are different from the novel ASVs in other samples. Since almost every sample for both twins contained *Klebsiella*, it was selected as an initial test case for novel strain differentiation.

### StrainID ASV sequence resolution enables differentiation of closely related *Klebsiella*

The analysis of the *Klebsiella* StrainID reads with DADA2 resulted in 54 ASVs that were present in each of the 24 infant fecal samples and three *Klebsiella* isolates K1, K7 and K8 (Figure 3A). *Klebsiella* genomes typically contain eight 16S-23S operons and the analysis of the amplicons from 3 pure cultures revealed the presence of a unique ASV fingerprint comprised of 7 different ASVs for each strain. The differences in relative abundance suggests that one ASV may be present as multiple copies.

**Fig 3.**
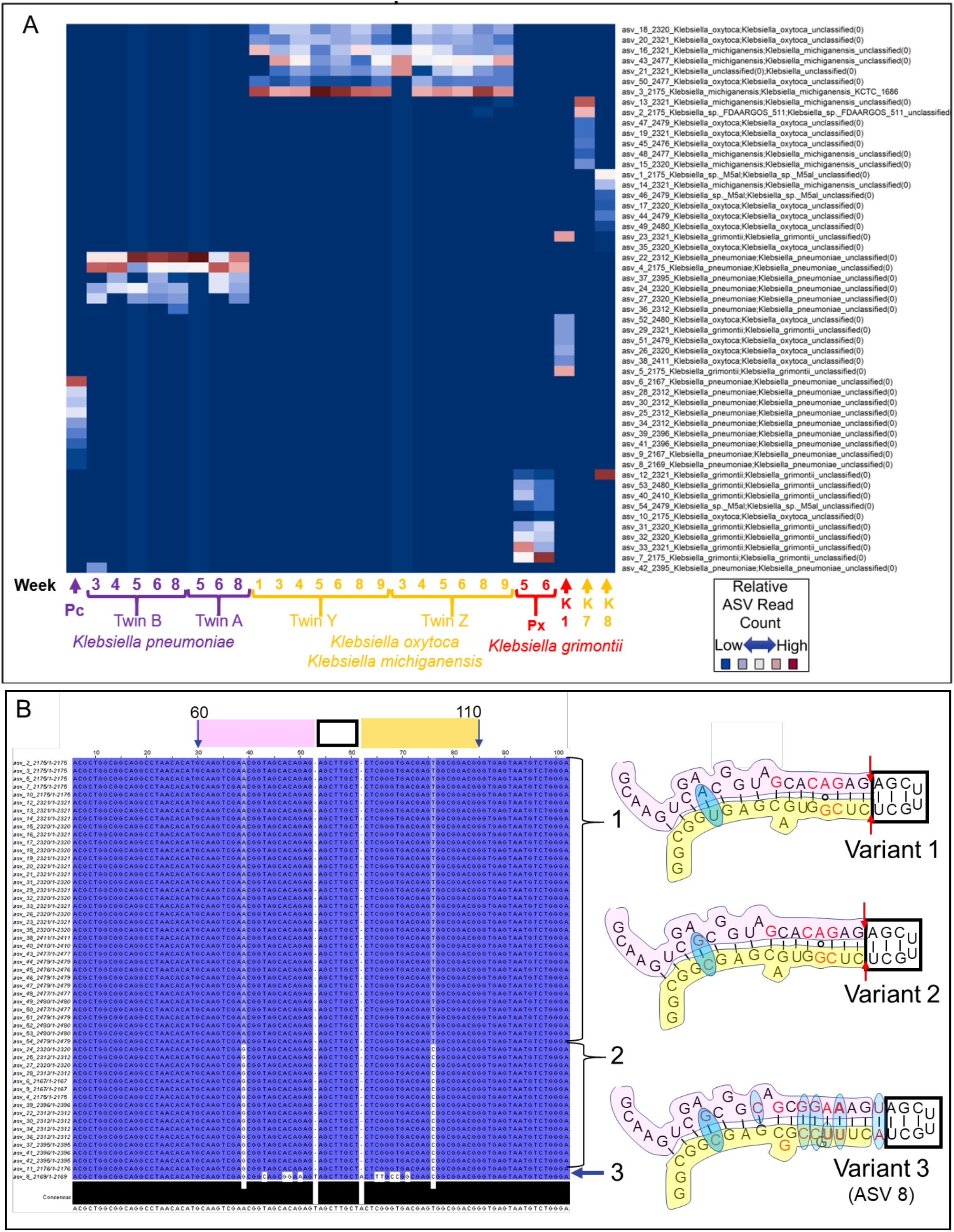
*Klebsiella* Amplicon Sequence Variant Fingerprints. **(A)** StrainID *Klebsiella* ASV Fingerprints. The figure shows a visual pattern of ∼2100-2500 base StrainID ASVs that make up a pattern, or ‘fingerprint’, in each sample, with the color indicating the relative read count assigned by DADA2 to each ASV in each sample. Samples sharing ASVs from the same *Klebsiella* genome will have similar or identical ASV fingerprint patterns. On the X-axis, longitudinal fecal samples IDs are indicated by brackets, numbers indicate the week since birth when the sample was obtained. Individual samples are indicted by arrows. ‘Pc’ and ‘Px’ indicate fecal sample ASVs from unrelated preterm infants classified as *K. pneumoniae* and *K. grimontii*, respectively. K1, K7 and K8 represent ASVs from *Klebsiella* isolates from unrelated preterm infants that developed necrotizing enterocolitis and were processed as single colonies. Reads classified by SBanalyzer as *Klebsiella* were analyzed using DADA2, resulting in the 54 *Klebsiella* ASVs listed on the y-axis. The ASV number is followed by the four-digit ASV length and the taxonomy assigned for that ASV by SBanalyzer. Each coordinate representing a ∼2500 base *Klebsiella* ASV in a sample was colored according to the relative number of reads assigned by DADA2 for that ASV in the sample, with dark blue indicating few reads and dark red indicating many reads. Possible species level taxonomic assignments for the fingerprint in each sample based on the best mapping results on the Y-axis are indicated by the label colors on the X-axis, purple for *K. pneumoniae*, yellow for *K. oxytoca/K. michiganensis*, red for *K. grimontii*. **(B)** *Klebsiella* Amplicon Sequence Variants in V1 maintain conserved stem-loop structure. All 54∼2500 base *Klebsiella* ASVs were aligned in Jalview (*28*), with each base numbered according to the alignment, and shaded by ‘Percentage Identity’. Dark blue variant bases match the most abundant variations, and white indicates a rare variant base. Bases 60-110 are indicated at the top of the alignment, part of the 16S V1 region containing a conserved stem-loop based on the *E. coli* 2-D structure (*39*). The 5’ stem, loop, and 3’ stem are marked above the alignment by pink, black and yellow boxes, respectively. Only three variant structures were identified at the V1 loop, shown at the right of the Figure. Brackets 1 and 2 indicate the ASVs with Variant 1 and 2, respectively. Arrow 3 indicates ASV 8, the only ASV with Variant 3 sequence. The pink, black, and yellow regions of the variant structures match the sequence in the alignment window. The red bases in the structures indicate *Klebsiella*-specific sequences common to these variants that differ from the *E. coli* structure. The red arrows in Variants 1 and 2 indicated a deletion from the *E. coli* and Variant 3 structures. The blue ovals indicate base variations between the aligned *Klebsiella* ASVs.

Analysis of the *Klebsiella* ASV fingerprint patterns revealed the long term colonization of premature infants by the same strain, and overall similarities between strains isolated between the twin pairs. Twins A/B shared one *Klebsiella* ASV pattern and Twins Y/Z shared another pattern suggesting that each pair was colonized by one strain and that these strains colonized each twin for at least 3 to 7 weeks. The ASVs in Twins A/B and infant ‘Pc’ were taxonomically classified as *Klebsiella pneumoniae*, although the ASVs were not 100% identical to strains in the Athena database. The ASV fingerprint in Twins A/B was distinct from the ASV fingerprint in sample ‘Pc’, indicating that Twins A/B and sample ‘Pc’ harbored distinct *K. pneumoniae* strains.

Twins Y/Z shared an ASV fingerprint over many weeks, indicative of the long-term colonization by a single strain. The ASVs in the fingerprint mapped most closely to either *K. oxytoca* or the closely related *K. michiganensis*. The Twin Y/Z ASVs were almost completely different from the ASVs for other *Klebsiella* samples, including samples from infant ‘Px’ and isolates K1, K7 and K8. *Klebsiella* ASVs from sample ‘Px’, from fecal samples taken in weeks 5 and 6 after birth from a non-twin preterm infant, contained ASVs that were classified as an unknown *Klebsiella grimontii* strain by mapping to the Athena database. *K. grimontii* is a species related to but distinct from *K. oxytoca* and *K. michiganensis* (*15*). The isolates K1, K7, K8, were cultured from fecal samples obtained from non-twin preterm infants with necrotizing enterocolitis (NEC) (*11*) and each contained a unique ASV pattern. *Klebsiella* ASV 13 and 2 were contained in more than one ASV fingerprint, they were present at high levels in isolate K7 and low levels in Twin Z (8 and 9 weeks). ASV 2 was observed in isolate K8 as well as sample ‘Px’. The low levels of ASVs 12, 13, and 2 seen in fecal samples could indicate a low-level copy number ASV that is far from the origin of replication in the *Klebsiella* in the fecal sample (*16*), a low level of co-infection with a second *Klebsiella*, or a low level of sample cross-contamination. However, the unique ASV fingerprints in the Y/Z twins, ‘Px’ and the NEC isolates suggesting they were 5 unique strains.

The ASV fingerprints convincingly demonstrate that StrainID ASVs can be used to differentiate closely related *Klebsiella* spp., and may have utility in following pathogenic strains in conjunction with other determinants of virulence in multiple individuals over time. For example, *Klebsiella* ASV fingerprints can be associated with complementary assays that determine whether they are toxin producing or not.

### ASVs Reflect Constraints of 2-Dimensional rRNA Structure, an Independent Confirmation of DADA2 ASV Accuracy at the Single Base Level

The Amplicon Sequence Variants produced from the StrainID sequence data were investigated further to explore the degree to which the sequence variations were underpinned by the biology of the 16S rRNA molecule. As an example, we depict the variation across all *Klebsiella* ASVs in the samples in a portion of the 16S gene V1 region in Figure 3B. The 2-D structures show that the variations in the V1 stem loop between bases 60 and 110 were not random, and reveal that the sequence variants fit with the hypothesis that they are evolutionarily constrained to maintain 16S function (*17*). This result is consistent with the observed variation in the ASVs, evidence that the DADA2-derived ASVs are an accurate representation of *Klebsiella* genomic variation and the result of PCR or sequencing artifacts. Although the ASVs originated in different individuals and time points, the overall variation at the V1 stem-loop was limited to three areas. The tip of the V1 loop, which was invariant in all 54 ASVs, is part of 16S helix 6, a region known as the ‘spur’, which is exposed to solution in the crystal structure and may be important in regulation of ribosome function (*18*). Variants 1 and 2 contained a double deletion indicted by red arrows, adjacent to the tip, which eliminated a base pair when compared to *E. coli*, but maintains an equal-length sequence on both sides of the stem. Variant 3 had a double insertion, similar to the A/U base pairing seen in *E. coli*, but in the opposite orientation. Variant 3 had a total of 13 bases that differ from Variant 1, and were represented by a single ASV, (ASV 8) of which 12 base changes maintained base pairing, and the thirteenth was an A/G substitution at a bulge that conserved a purine at that site. The *Klebsiella* ASV bases in red text that differ from the *E. coli* reference structure also maintained stem-loop structure in a similar manner. For each V1 region inferred for the 54 *Klebsiella* ASVs, overlaying the base-level variation observed in this 100-base window revealed that the ASVs generated by DADA2 were consistent with maintaining 16S rRNA structure, because the base changes fit a pattern where the base pairing in the V1 stem loop between bases 60-100 was preserved. The observation that the base-level variation in the ASVs is consistent with maintenance of the 2-dimensional structure of the 16S rRNA molecule is strong evidence that the ASVs generated from the sequence data by DADA2 are representative of the true genomic variation of the bacterial genes across all ∼2,500 bases of the StrainID amplicon.

Investigation of *Klebsiella* species demonstrated that long amplicons can be used to differentiate novel, closely related species and strains in a sample set. Similar methods were used to determine the relatedness of *Enterobacter* and *E. coli* in an expanded infant fecal microbiome sample set to demonstrate that the methods apply to other important bacteria in the infant microbiome.

### Enterobacter ASV Fingerprints Enable ID and Tracking of Strains Across Samples

In Figure 4A, *Enterobacter* ASVs were inferred from Twin A, Twin B, and Infant 961 samples, all of which contained significant quantities of *Enterobacter* at multiple time points. The resulting ASVs were mapped to NCBI using BLAST, which resulted in six 99%+, but non-identical, matches to a variety of strains for each sample. *Enterobacter* typically contain eight 16S-23S regions. However, a scan of *Enterobacter* strains in the Athena database revealed that one or more regions may be an exact duplicate within a genome, resulting in fewer than 8 amplicons per genome (data not shown). Since *Enterobacter* genomes are expected to produce 6-8 unique StrainID amplicons that would map to a single strain, the variety of imperfect mapping results are an indication that the strains present in the three infant microbiomes were from previously unsequenced strains. For each individual, the same sets of ASVs persisted over the weeks sampled, evidence that the novel variants are not artifacts. For example, PCR or other sample-dependent 2000+ base ASV artifacts would not be expected to be present in multiple time points and/or individuals. Reproducibility of ASVs within a sample is shown by comparison of Twin B technical replicates 6 and 6r, which yielded identical ASVs in similar relative proportions. Twins A and B contained the same overlapping set of 6 ASVs, starting one week apart, and continuing through week 8. These twins, admitted to the same NICU at the same time, were apparently colonized a week apart by the same *Enterobacter cloacae*. In contrast to the twins, Infant 961 contained ASVs that mapped primarily a variety of regions from *Enterobacter hormaechei* strains. Antibiotic treatment at week 17 resulted in the loss of all *Enterobacter* ASVs, so that time point is not represented. However, the same ASVs reappeared by week 18, indicating that either *Enterobacter* was reduced to undetectable levels, but not completely eliminated from the gut, or re-infection of the same strain occurred.

**Fig 4.**
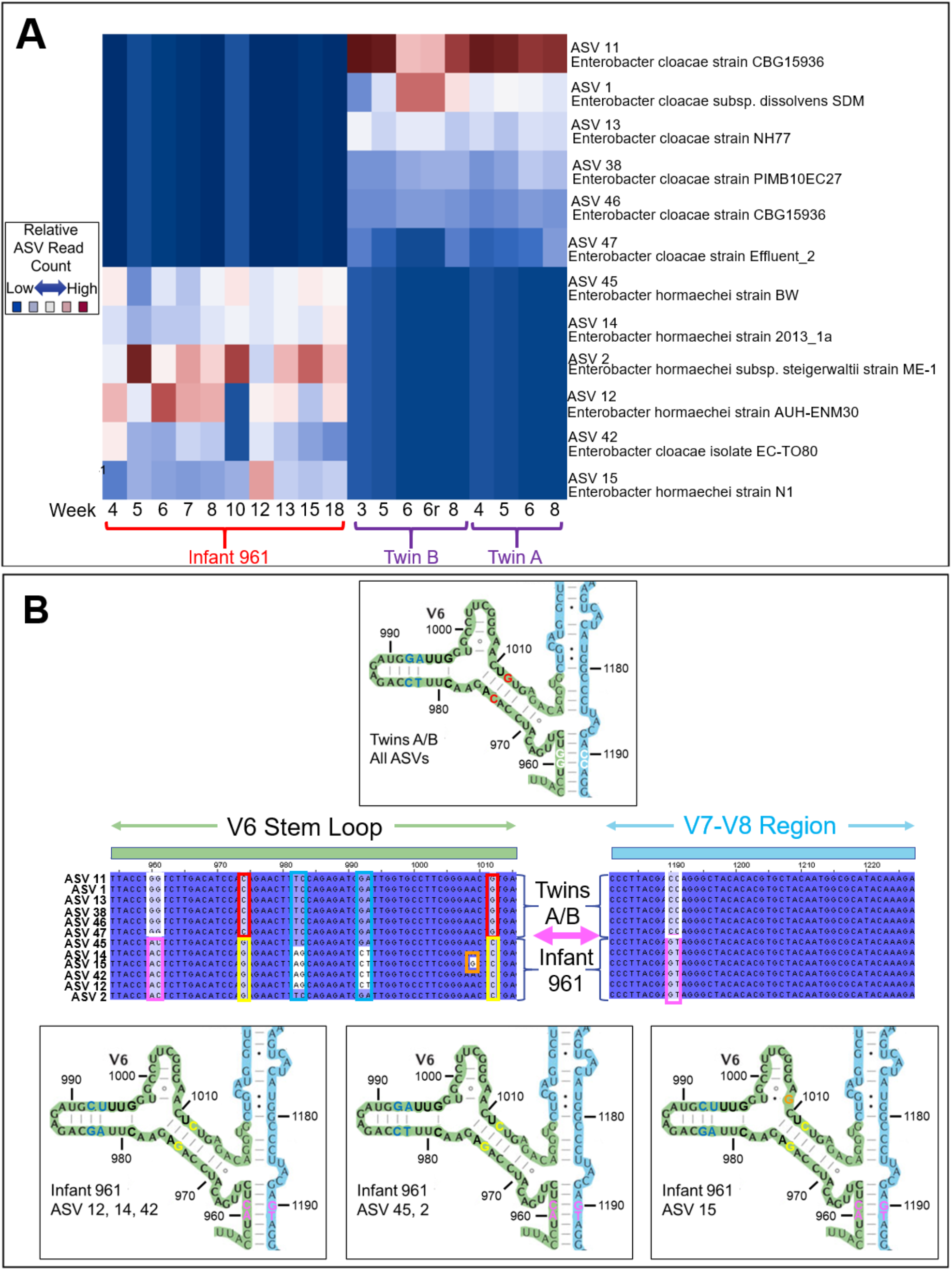
*Enterobacter* Amplicon Sequence Variant Fingerprints. **(A)** *Enterobacter* ASV fingerprints inferred from infant fecal samples. ASVs were inferred from longitudinal samples at the indicated weeks from reads classified as *Enterobacter* by SBanalyzer from Twins A and B, and an additional individual Infant 961, indicated by brackets. All ASVs were mapped to NCBI and the closest matching taxonomies were selected, and displayed on the Y-axis to the right of the figure. The relative number of each of the ASVs in each sample are indicated by the shading, where dark red indicates more reads, and blue indicates fewer. Points 6 and 6r in Twin B are technical repeats of the week 6 fecal sample. **(B)** Enterobacter ASV Mapping to 2-D 16S rRNA structure. The twelve *Enterobacter* ASVs inferred from Twin A, Twin B and Infant 961 using DADA2 were aligned against each other in Jalview (*28*), with each base numbered according to the alignment, and shaded by ‘Percentage Identity’. Dark blue variant bases match the most abundant variations, and white indicates a rare variant base. The stem-loop rRNA structures are based on *E. coli* (*39*). The selected sequences shown in the center of the Figure were mapped to the 16S gene structure from bases 955-1015 (the V6 stem-loop, green box above sequence) and 1180-1230 (light blue box above sequence, region between V7 and V8). Identical sequences across all ASVs are shown in black text with dark blue highlight. Variant sequences are shaded and boxed with different colors, indicating the text color of the variant bases in the corresponding structures. The structure above the alignments represents all Twin A/B ASV structures, which are identical in the regions shown. There are three variant structures in this sequence shared within the Infant 961 samples, shown below the alignments. The variations in the V7-V8 region from 1190-1191 form a base paired structure with the V6 region ∼200 bases away at bases 960-961.

In Figure 4B, *Enterobacter* variants inferred from the different samples were aligned against each other and mapped back to the 2-D 16S rRNA structure to determine whether the variations between ASVs maintained the base pairing and other expected structural constraints. The 16S-V6 region was selected because of the long-distance pairing near base 960 with variations 230 bases away between the V7-V8 regions near base 1190. This long range intramolecular interaction is a clear demonstration of the ability of DADA2 to infer accurate single base calling for these ASVs. The portion of the *Enterobacter* ASVs that mapped to the 2-D structure in the V6 region and the structurally associated sequence between the V7 and V8 demonstrate that the variants inferred by DADA2 correspond to predictable combinations of sequence variations dictated by 16S rRNA structure. The ASVs in the twin samples shared 8 distinct differences in the V6 loop that contrasted with the ASVs in Infant 961. This number of differences serve as clear differentiators between the regions obtained for the two samples, mapping most closely to *Enterobacter cloace* for the Twins A/B and *Enterobacter hormaechei* for Infant 961. As in this example, it is expected that regions within a genome would generally be more similar than regions between genomes. However, there were 4 shared variations at bases 962-963 for Twins A/B and two of the ASVs from Infant 961, ASV 45 and ASV 2. Of note is the single base change at position 1,009 that helped to differentiate ASV 15, where an A/U base pair is substituted by a G/U base pair, maintaining base pairing in the stem. Because the G/U base change in ASV 15 was maintained across 18 weeks for ten different time points, and the base change maintained the 16S base paired structure, it is likely that this single base change alone is a true variant that can serve as a meaningful differentiator separating ASVs.

### *E. coli* ASV Fingerprints Enable ID and Tracking of Strains Across Samples

*E. coli* commonly occurs in human microbiome samples, and this species includes a variety of strains that can vary in impact from benign to pathogenic. DADA2 was used to infer ASVs from reads SBanalyzer identified as *E. coli* from the four twin samples A/B and Y/Z, and the resulting fingerprints were compared across all time points in Figure 5A.

**Fig 5.**
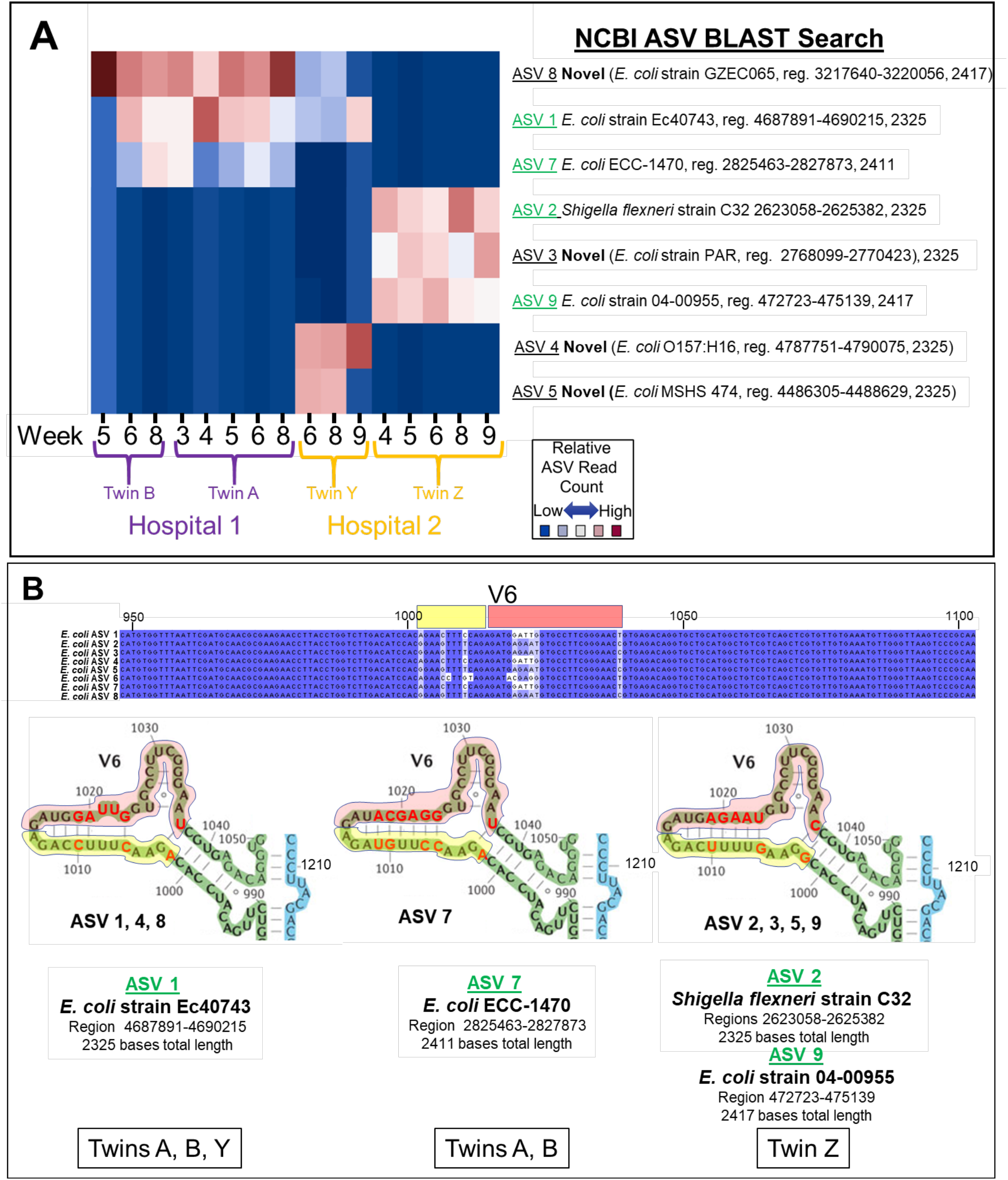
*E. coli* Amplicon Sequence Variant Fingerprints. **(A)** *E. coli* ASV fingerprints in twin fecal microbiome samples. ASVs were inferred from longitudinal samples at the indicated weeks from reads classified as *E. coli* by SBanalyzer from Twins A/B and Twins Y/Z. Twins A/B were admitted simultaneously to Hospital 1, and Twins Y/Z were admitted simultaneously to Hospital 2, as indicated. The 8 ASVs obtained from the samples using DADA2 were mapped to NCBI, the closest matching taxonomies were selected, and displayed to the right of the Figure. ASVs in green text indicate perfect matches to the indicated NCBI region. Bold text ‘Novel’ indicates an ASV that did not match perfectly to any sequence in NCBI, but was present in multiple samples. The twin ID and the week the sample was collected is indicated below the figure. The relative number of each of the ASVs in each sample are indicated by the shading, where dark red indicates more reads, and blue indicates fewer. **(B)** *E. coli* ASV structure at the V6 stem loop. *Escherichia coli* ASVs were inferred from Twins A/B and Twins Y/Z using DADA2, and the eight resulting ∼2300-2400 base 16S-ITS-23S ASVs were aligned against each other in Jalview (*28*). A portion of the alignment from bases 948-1103 containing the V6 region is shown, with identical sequences across all ASVs in black text with dark blue highlight, and ASV specific variations shaded in lighter colors. The yellow and red boxes above the alignment indicate the V6 stem-loop sequences shaded in the structures pictured immediately beneath the alignment in the middle of the figure. The 2-D structures of the aligned V6 region of each ASV sequence was mapped to the *E. coli* 16S gene structure (*39*) from bases 988-1050, bases were numbered as per the alignment. Variant sequences from the alignment are depicted in bold red text. Yellow and red shading in the structures corresponds to the yellow and red boxes above the aligned sequences.The ASVs numbers containing the exact stem-loop V6 sequence are listed below each structure. ASVs with NCBI BLAST searches that produced perfect matches to the entire V6 sequence are listed immediately below the structure in bold green underlined text. The NCBI BLAST strain ID, genomic region, and ASV length for the perfect full-length ASV match containing the V6 sequence are listed immediately below the bold green text. The individual twin samples containing ASVs that include each V6 sequence are listed at the bottom of the figure in the outlined boxes.

The *E. coli* ASV fingerprint results reveals similar findings as for *Klebsiella* and *Enterobacter*, where the one ASV fingerprint was present in Twins A/B and that it was distinct from the ASV fingerprints identified in Twins Y/Z. Twin A was colonized at week 3, whereas Twin B was colonized two weeks later. However, Twins Y/Z had two completely different *E. coli* ASV fingerprints. Twin Y samples contained unique ASV 4 and ASV 5, whereas Twin Z samples contained ASVs 2, 3, and 9. The fact that no ASVs were shared by Twin Y and Twin Z is evidence that they were colonized by different *E. coli* strains, even though they were admitted to the same NICU at the same time.

The sequences of the eight *E. coli* ASVs obtained from the 16 longitudinal samples from 4 twins were compared to the *E. coli* 16S rRNA 2-D structure, the sequences in the V6 region are shown in Figure 5B. NCBI BLAST searches of *E. coli* ASVs revealed that the *E. coli* in the twin samples contained a combination of existing and novel *E. coli* 16S-23S regions. Furthermore, the alignments demonstrated that the novel ASVs contain sequences that are combinations of known structures that were identified in unrelated sequencing projects that combine to create the novel ASVs found in this study. The fact that each of the novel ASVs were identified at multiple time points, and that ASV 7 was identified in two individuals, and ASV 8 and ASV 1 were identified in three separate individuals as subsets of a unique group of ASVs (and therefore likely a unique, previously unsequenced *E. coli*) demonstrates that the novel sequences are unlikely to be artifacts. It is notable that four of the eight ASVs were perfect matches over the 2,300-2,400 base length of the StrainID amplicon. The sum of these observations supports the hypothesis that novel ASVs can be a combination sequences/structures shared with other bacterial 16S sequences, and that DADA2 is capable of inferring very large 16S-ITS-23S ASVs with single base resolution.

Interestingly, Twin Y shared *E. coli* ASV 1 and ASV 8 with Twins A/B. A search of the Athena database showed that there are a number of 16S-23S regions shared across *E. coli* strains, but there are few sequenced strains that share all regions (Supplementary Fig. S2). The fact that the Twins A/B and Twin Y share only a subset of ASVs, and they were admitted to different hospitals at different times indicate that they acquired different *E. coli* strains that share a subset of ASVs. The illustration *in silico* and in infant fecal samples is an indication that while different strains of *E. coli* can share a subset of ASVs, the variation in combination of the ASVs (ASV fingerprint) provides sufficient resolution to enable strain-level tracking of bacterial transmission for closely related bacteria in hospital environments.

### *Klebsiella* Reference Database Genome Taxonomy Investigation

Next, we wanted to assess the accuracy of the species identification using the Athena database. Identification of bacterial taxonomies using either sequencing read data or ASVs requires that data be accurately mapped to a database containing correct taxonomies. The Athena database was created from well-sequenced genomes in the public domain, but it has been documented that there are many mis-identified genomes in GenBank, and even though best practices have been suggested (*19*) they are not always followed. Additionally, existing entries in a database may not be updated as nomenclature is refined over time, so closely related species that were described at different times can sometimes be assigned different taxonomies based on the best practices upon publication (*20*). To address the potential for ‘mis-assignment’ of taxonomies for sequenced genomes, three methods were used to explore the ‘correctness’ of *Klebsiella* taxonomies within the Athena database, two based on 16S and one based on whole genome sequences. There are a total of 458 *Klebsiella* genomes with 3,635 regions in the Athena database. In Figure 6, a randomly selected total of 1387 *Klebsiella* full-length16S genes and 16S-23S StrainID amplicons were extracted from the Athena database, and phylogenetic trees were created to assess how well the taxonomies matched the sequence variability. Visualization of the taxonomic tree for the 16S rRNA gene alone indicated that the different *Klebsiella* species mostly clustered as expected, with *K. oxytoca* and *K. michiganensis* somewhat interleaved, but mostly separated from *K. aerogenes* and *K. pneumoniae*. The increased diversity contributed by the longer StrainID amplicon resulted in dispersal of regions that sorted together in the 16S tree into multiple groups. This demonstrates that the longer StrainID amplicon enables improved differentiation of amplicons from similar organisms, but dramatically increased sequence diversity introduced by inclusion of the intergenic spacer region may be a larger driver of similarity than the differences based on the relatedness of the 16S genes. For example, the eight 16S and 23S genes in most *Klebsiella* genomes contain spacer regions with either no tRNA genes, a single tRNA Glu gene, or a both tRNA Ile and tRNA Ala, introducing changes to both sequence and amplicon length that may drive 16S-23S region-specific rather than species-specific clustering in the StrainID taxonomic tree. This would not present an issue when assigning taxonomies to known genomes, but for unknown genomes the possibility of novel content in the ITS region should be considered when considering putative taxonomic assignments, along with the possibility that the closest mapping 16S region may not be assigned because of variability in the intergenic spacer region.

**Fig 6.**
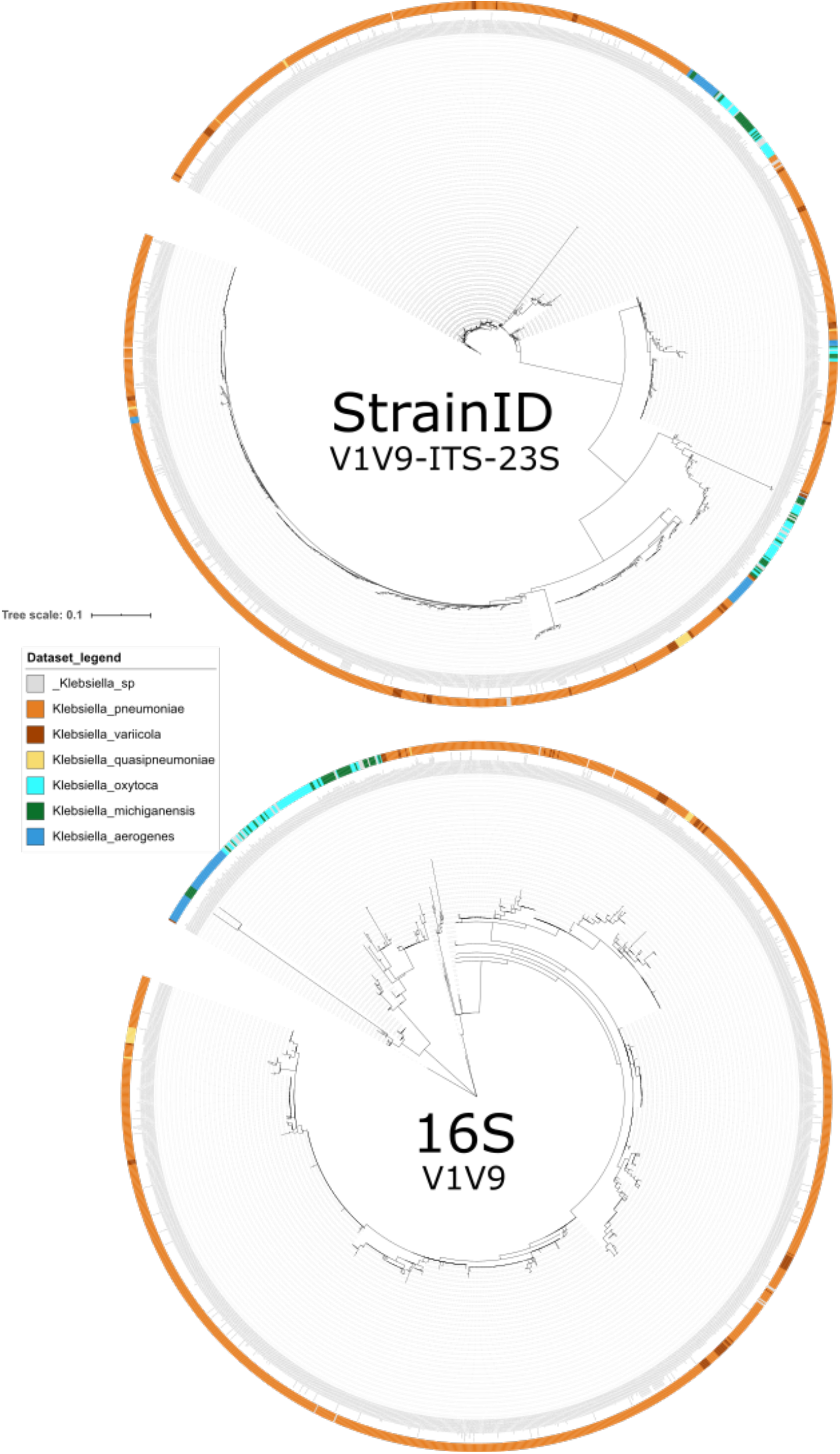
Taxonomic Tree of *Klebsiella* 16S and 16S-23S StrainID Amplicon Sequences. Amplicon sequences from Athena database were extracted using the 27f StrainID forward and either the 1492r (16S V1V9) or StrainID Reverse (V1V9-ITS-23S) sequences from Materials and Methods. The sequences were aligned in Geneious Prime version 2020.1 using Clustal Omega(*29*) and the phylogeny inferred using RAxML(*30*). The tree was annotated in iTOL(*31*)

The Athena database is primarily constructed from well sequenced genomes that are publicly available, where the associated taxonomies were assigned by the investigators at the time the sequences were uploaded to the database. As a result, taxonomies may not be updated as conventions change, and may become outdated, which should be considered when proposing taxonomic assignments to newly discovered bacteria with unique ASV fingerprints. To assess taxonomic designation accuracy using existing assignments to publicly available genomes, ten genomes labeled *Klebsiella oxytoca* with 16S-23S regions represented in the Athena database were compared with 11,300 bacterial strains from the TYGS database using both the 16S sequences and the full genomes (Supplementary Fig. S3). The whole genome phylogram revealed that two of the “*Klebsiella oxytoca*” Athena strains were closest to *K. grimontii*, five were closest to *K. michiganensis*, and only three were most closely related to *K. oxytoca*. The relationships inferred by the 16S rRNA gene phylogram were similar, where the three most closely related to *K. oxytoca NCTC 13727* were clustered together, but four of the five most closely related to *K. michiganensis* sorted differently. The two *K. grimontii* related strains from Athena sorted together as with the whole genome results, but far from the *K. grimontii* from the TYGS database. The differences between the whole genome and 16S methods indicate that the 16S phylogeny may not always reflect the whole genome phylogeny and that the taxonomic names in the databases are not always correct. A database such as Athena, built primarily from genomes obtained from the public databases, is likely to contain sequences that were assigned taxonomies that do not match whole genome comparisons that would have been assigned more recently. Although the ASVs inferred from the sequence data may reveal previously unsequenced 16S-23S regions from novel bacterial strains, the differential sorting of whole genome and 16S methods indicates that 16S phylogenies alone may not always be sufficient to accurately assign optimal species or strain level taxonomies to novel genomes.

We further explored the discrepancy of the 16S rRNA and whole genome phylogenies by examining the individual regions from each of the 10 *Klebsiella* strains from the phylogram analysis. All of the 16S rRNA gene copies were mapped individually to determine the taxonomy assigned to each individual 16S rRNA gene (Supplementary Fig. S4). As expected, the genomes labeled as either *K. michiganensis* and *K. oxytoca* by whole genome TYGS phylogeny contained regions that all mapped to the corresponding organism. Interestingly, *K. grimontii* genomes contain a blend of regions common to both *K. michiganensis* and *K. oxytoca*. This result indicates that although the taxonomy of a genome may not be up to date or the 16S rRNA gene phylogeny may not reflect the taxonomy, an understanding of the 16S gene combinations for organisms of interest provides an additional method for discrimination of closely related taxonomies.

### Amplicon Length Correlates with Utility for Taxonomic Discrimination

The length and region of the rRNA operon dramatically affects the taxonomic resolution obtained. The widely used ∼250 base V4 region of the 16S rRNA gene usually allows the identification of the bacteria to the family or genus level. A V1-V9 ∼1,500 base 16S rRNA gene amplicon can frequently yield taxonomic specificity at the genus or species level (*6*), but closely related bacterial strains may have identical 16S genes. In order to compare the ability of different amplicons to uniquely identify a set of known genomes, an *in silico* comparison of 458 *Klebsiella*, 187 *E. coli*, and 109 *Enterobacter* was performed for amplicons covering the V4, V1V3, V1V9, and the 16S-23S StrainID regions (Supplementary Fig. S2) extracted from the newly created Athena database. A genome ID was considered unique if it contained either a unique amplicon, or a unique combination of amplicons. The trends for all three bacterial taxonomies demonstrate that for a given genus and species, most V4 amplicons are identical, whereas most StrainID amplicons are unique. For example, each of the 458 *Klebsiella* genome ID entries in the Athena database contains up to 8 individual 16S-23S amplicons, so all eight regions from each *Klebsiella* in the database could be compared to the 8 regions from all others, to determine how many *Klebsiella* had a unique sequence or unique combination of amplicons that enable differentiation from all other genomes in the database. Relative abundance of identical amplicon duplicates (if they occurred) was not considered a difference for the purposes of this analysis. Figure S2 shows that only 63, or 14%, of the *Klebsiella* genomes could be uniquely identified using the ∼300 base V4 region amplicon. The ∼526 base V1-V3 amplicon sequences enabled unique identification of approximately half of the genomes in the Athena database, the others had one or more exact duplicate profiles. The full ∼1500 base 16S gene V1-V9 amplicon enabled unique identification of 285 genomes, whereas and StrainID amplicon enabled unique differentiation of 344 of the 458 genomes, 75%. This number of uniquely identifiable genomes may be an underestimate, because the Athena database was constructed from well-sequenced genomes, but without regard to whether the genomes are exact repeats of previously sequenced clones, so there may be duplicate sequences of identical strains in the database.

## Discussion

One of the major challenges of microbiome studies is the limited taxonomic resolution that the short stretches obtained by traditional 16S rRNA amplicon sequencing provide. Amplification and sequencing of the 2,100 to 2,500 bp 16S-23S StrainID portion of the rRNA operon, and comparing the sequences to the genome-based Athena database allows the identification of bacterial species and known strains. DADA2 can be used to reveal strains with unique amplicons or unique amplicon combinations that generate correspondingly unique ASV fingerprints. We tested this novel approach in premature infants that harbor a simple microbiome by analyzing longitudinal time series of fecal microbiome profiles from individual infants and pairs of twins.

Multiple strains of *Klebsiella* spp., *E. coli*, and *Enterobacter* spp. were detected in fecal samples of the five infants in this study. Amplicon sequence variant analysis using DADA2 indicated that individual samples contained 16S-ITS-23S gene combinations or ASV fingerprints that were not present in the Athena database or NCBI, suggesting that they represented either unknown strains or strains without closed genomes. The variable ASV sequences inferred using DADA2 could be assigned to the 2-D structure of the 16S molecule, demonstrating that the base changes were not random, that they tended to vary such that the 2-D hairpin loop structure of known variable regions were maintained. Single base differences between ASVs inferred by DADA2 were supported by both structural data and by identifying identical sequences in the NCBI database. In some cases, the inferred ASVs were 100% matches to sequences in the NCBI database for a particular species, although there was no case where all of the ASVs matched all of the 16S-23S regions for a known genome. The ASVs created from the StrainID sequencing data from infant fecal samples, including multiple sequential time points, duplicate samples, and twin pairs, all provided independent confirmation of ASV sequences representing the presence of strains of previously unsequenced *Klebsiella* spp., *E. coli*, and *Enterobacter* spp. that could be identified to the species level.

The increased resolution provided by the combination of the StrainID amplicon, the Athena database, and the DADA2 ASVs were strongly supported by the experimental design which included longitudinal time series of samples obtained over consecutive weeks, enabling independent confirmation of novel ASVs over time in independent sampling events. Twin samples from individuals co-admitted enabled determination of whether individuals carried a single strain over time and determine if two individuals shared a common strain. The unique ASV fingerprints from the stool samples from twin pair A/B, indicate that they shared one *Enterobacter* cloacae strain, *Klebsiella pneumoniae* strain, and one *E. coli* strain across multiple time points. Twin pair Y/Z carried the same *K. grimontii* strain, two distinct *E. coli* strains and no *Enterobacter* strain. In addition, DADA2 inferred ASVs for *Klebsiella* and *Enterobacter* from non-twin infant fecal samples that were completely different. Interestingly, although Twins A and B had an exactly matching *E. coli* ASV group profile, Twin Y and Twin Z did not share any common *E. coli* ASVs. This result supports the conclusion that although Twins Y and Z were admitted at the same time in the same NICU, they were colonized by different *E. coli* strains. It was clear that the *E. coli* in Twin Y shared at least 2 regions with Twins A/B, consistent with the *in silico* results of the Athena database comparison of *E. coli* sequences, which showed that different *E. coli* sometimes share subsets of StrainID ASVs. The *E. coli* results demonstrate that although there may be examples of strains that share one or more identical ASVs, the total complement of StrainID amplicons or the ASV fingerprint is useful in differentiating closely related bacteria, revealing colonization patterns and temporal microbiome dynamics.

Although the StrainID amplicon generally enables higher resolution taxonomic classification than the 16S gene alone, there are a number of limitations that should be considered. For example, even if ASVs obtained from a sample match a known strain, it does not necessarily confirm the genome in the database is an identical match, because closely related bacteria, including as-yet unsequenced strains, may share an identical set of StrainID amplicon fingerprints while harboring important differences elsewhere in the genome. The fingerprints for the different *Klebsiella* spp., *Enterobacter* spp., and *E. coli* discussed in this report were different from the sequence combinations in any sequenced genomes, indicating that they are novel strains. In fact, the ASVs for those three bacterial strains were unique, with the exception of a subset of *E. coli* ASVs. While unique ASV fingerprints indicate real genomic differences, those differences may be limited to a few bases or even a single base in an amplicon, and should be considered in context, for example, whether the differences are consistent across multiple individuals or time points. Ideally, experiments should be designed with technical repeats or longitudinal sampling such that conclusions are not based on variants identified in a single sample. Yet another important consideration is that not all ASVs will necessarily represented at ratios corresponding to their relative genomic copy number, because the distance of ribosomal operons relative to the origin of replication can drive increases or decreases of relative copy number. For example, during chromosome replication, genes close to the origin of replication will be present at higher copy number than those closer to the terminator, especially if the cells were growing rapidly at the time the sample was acquired (*16*). As a result, ASVs from known strains may be missed if the sequences are under-represented due to growth and environmental effects. Genomic structure is another important consideration for the StrainID amplicon, there are species important in human gastric samples (e.g., *Helicobacter*) or environmental samples (e.g., certain Planctomycetes) where the 16S and 23S genes are thousands of bases apart, so no amplicon will be generated from these bacteria. In the cases where genomes without proximal 16S-23S genes are prevalent, a lower resolution 16S amplicon such as V1-V9 region may be preferred.

StrainID fingerprints of *Klebsiella* spp., *Enterobacter* spp., and *E. coli* were unique, even though these taxa are well-represented in the Athena and NCBI databases. The fact that only novel bacterial strains were identified is an indication that novel strains may be commonly encountered in fecal microbiomes. ASV fingerprints can be used to identify novel organisms, but the resulting taxonomic classification of the novel ASVs is only as accurate as the associated taxonomies in the database. Our investigation into *Klebsiella* taxonomic assignments using publicly available genomes reveals that it is important to consider whether the taxonomies that are attached to the best sequence matches in a database were correctly assigned, and whether any important updates to the classifications were made after the sequences were posted. For example, *K. grimontii* was described in 2018 as separate from *K. oxytoca* and *K. michiganensis*. Furthermore, *K. grimontii* strains contain subgroups of ASVs, some of which map most closely to *K. oxytoca*, and some to *K. michiganensis*. Care should be taken in the assignment of species and strain level classification of newly discovered bacteria so that new assignments are as correct as possible.

In summary, the StrainID rRNA amplicon provides a higher level of taxonomic information than the full 16S gene, and can therefore provide a higher-resolution taxonomic picture of the overall microbiome. *Klebsiella, Enterobacter*, and *E. coli* were used as examples to demonstrate the ability to differentiate closely related taxonomies of specific early microbiome colonizers in infants confined to the NICU. Similar methods can be used to obtain high-resolution ASVs for other target bacteria using the StrainID amplicon. The assay provided a high-resolution longitudinal view of the colonization of the gut in premature infants, which may be useful for tracking the results of therapeutic interventions as well as opening a window into understanding how commensals are established in the human microbiome after birth. It may be possible to use the assay to identify improvements to treatment that establish important commensals earlier, for improved health outcomes over short-term hospital stays and long-term development. The StrainID amplicon provides a rapid, practical, high throughput, and high-resolution method of identifying and tracking known and unknown pathogens and commensal bacteria in the complex environment of the microbiome.

## Materials and Methods

### Experimental Design

Longitudinal fecal samples from seven infants, including two sets of premature twins, born with gestational ages close to 30 weeks and cared for in two different NICUs, were collected each week from admission to the NICU to discharge. Metadata for each infant was collected concurrently, including antibiotic treatment, any diagnosed infections or dysbiosis, and nutritional sources. Bacterial content of infant fecal material was analyzed using the ∼2,500 base StrainID amplicon to determine whether strain-specific amplicon sequences could be used to follow the development of microbiomes in the infants at high resolution. Specifically, the objective was to assess whether StrainID amplicon could be used to follow strains of known and unknown pathogens and commensals over time and across individuals during microbiome development, infection, and treatment. The design also included *Klebsiella* isolates obtained from infants previously treated in the NICU as strain level controls for the analysis, to determine whether StrainID amplicon could be used to differentiate the isolates, and whether signal from the same isolates could be identified in the study.

### Fecal sample isolation and PCR

Preterm infant fecal samples were obtained from subjects cared for at two affiliated NICUs in Hartford and Farmington, CT. Infants were enrolled during 2018 as part of an ongoing neonatal microbiome study approved by the Institutional Review Board at Connecticut Children’s Medical Center. Infant fecal samples were collected by trained bedside nurses on an approximate weekly basis beginning with the first bowel movement until discharge. Samples were collected using sterile, disposable spatulas during diaper changes, placed into sterile containers, and immediately frozen at −80°C until processing. Approximately 1-5 mg of fecal material was used as input into the StrainID kit (Shoreline Biome, cat# StrainID Set A [Barcodes 1-96]). Fecal sample DNA was isolated as per manufacturer’s instructions. Briefly, 50 µl of reconstituted lysis reagent was added to each sample. Subsequently, 50 µl of 0.4 M KOH solution was added and the samples were heated to 95°C for 5 minutes to lyse the cells. The plate was spun briefly to pellet the fecal debris, and 50 µl of the supernatant was transferred to a clean plate. 50 µl of DNA Purification Beads were added, the DNA was allowed to bind, and the pellets were washed with 70% ethanol. DNA was eluted in TE, and 10 µl eluted DNA was transferred to the corresponding well in the PCR plate. 2x PCR mix was added, and PCR was performed as per instructions to amplify and barcode each sample. Samples were pooled, purified via spin column (Qiagen MinElute, cat # 28004).

### Individual *Klebsiella oxytoca* isolates

*Klebsiella oxytoca* fecal isolates were from non-related preterm infants that developed necrotizing enterocolitis (*11*). To obtain individual isolates, a loopful of fecal material was inoculated into 5 ml of pre-warmed Lysis Broth (Lennox) broth with vitamin supplementation (ATCC MD-VS). After overnight growth at 37° C, 1 ml of culture was serially-diluted in PBS and plated onto *m*-hydroxybenzoic agar medium (*21*). After 24 - 36 hours of incubation at 37° C, individual colonies were screened by colony PCR for *pehX*, a genetic marker specific for *K. oxytoca* (*22*). Purified genomic DNA from *K. oxytoca* ATCC strain 13182 was used as a control. The PCR reaction was performed using iproof high fidelity PCR Kit (Bio-Rad) with the primers: Forward – 5’-GATACGGAGTATGCCTTTACGGTG-3’, and Reverse – 5’-TAGCCTTTATCAAGCGGATACTGG-3’. Post PCR identification, individual colonies were streaked onto *Klebsiella* select agar (Sigma Aldrich, St. Louis, MO) and grown overnight. Isolated colonies were then processed for biotyping using MicroScan WalkAway 40 (Beckman Coulter, Brea, CA), and antibiotic susceptibilities were determined in the UConn Health Clinical Microbiology Laboratory. For whole genome sequencing, genomic DNA was extracted using MasterPure DNA Purification Kit (Lucigen) and used for Nextera XT (Illumina) based amplicon library preparation and sequencing on Illumina platform at the Microbial Analysis, Resources, and Services (MARS) facility at the University of Connecticut.

### StrainID Amplicon and 16S gene sequences

The StrainID amplicon spans the full 16S and partial 23S rRNA genes. Forward and reverse primers were synthesized as a 3-part sequence as follows: 5’-Adaptor-Barcode-TargetSpecificPrimer-3’. The 16S Adaptor sequence is 5’-GGTTATGCGGTTCACTGC-3’. All barcodes were selected from the list of 384 PacBio recommended barcodes (https://www.pacb.com/products-and-services/analytical-software/multiplexing/). The target-specific forward primer sequences used are a pool of primers with the sequences 5’-AGRRTTYGATYHTDGYTYAG-3’. The reverse primer had a similar 5’-Adaptor-Barcode-TargetSpecificPrimer-3’ structure, where the Adaptor sequence is 5’-

CGTCACTTGGCGTATTGG-3’, and the target specific sequences are a pool with the sequences 5’-AGTACYRHRARGGAANGR-3’. The forward primer is located at the start of the 16S gene, whereas the 23S primer site is located about 600 bases inside the gene. Both primer sites were used as a starting point to identify additional primer site variants that exist in bacteria found in the following databases: Shoreline Biome Athena, SILVA 16S and 23S (*23*), and Riken (https://metasystems.riken.jp/grd/download.html).

For the 16S rRNA gene sequences extracted from the databases for *in silico* comparisons, the reverse primer used was based on the 1492r primer: 5’-TASVGHTACCTTGTTACCGACTT-3’.

### DNA sequencing

Amplicon library were created using the SMRTbell Express Template Prep Kit 2.0 (PacBio, cat# 100-938-900) as per manufacturer’s instructions. Library was sequenced on a Sequel1 (Pacific Biosciences) at the University of Delaware, Delaware Biotechnology Institute Sequencing & Genotyping Center, Newark, DE. 320,054 ccs reads were produced using default settings.

### Taxonomic Assignment of Reads

SBanalyzer 2.4 (https://www.shorelinebiome.com/sbanalyzer-software/) was used to map ccs reads to the Athena database and assign taxonomic identification to all reads. SBanalyzer produces a summary ‘.csv’ file with bacterial taxonomic ID and corresponding read count for all samples that can be manipulated in Excel, LibreOffice, or other spreadsheet program, as well as a ‘.taxonomy’ file (Supplementary Data File S3) with the taxonomic assignment and a ‘.groups’ file (Supplementary Data File S4) with the sample assignment for each read. SBanalyzer is a graphical user interface-based pipeline that encapsulates custom algorithms and external calls to version 1.40.5 of ‘mothur’ (*24*). However, the ∼2,500 bp amplicon reads required significant changes to the default ‘mothur’ methods for both demultiplexing and mapping for optimal results. Demultiplexing of barcodes and assignment of reads to samples employed custom code optimized for 16-base PacBio barcodes combined with the dual-unique barcode structure of the primers, which was able to assign 97-99% of ccs reads to a sample. In addition, mothur was customized to use BLAST-plus (*25*) to map the ∼2,500 bp amplicon reads, since the standard mothur algorithms and settings failed to correctly assign long amplicon taxonomies.

### Athena Database

The Athena database is an integrated part of the SBanalyzer pipeline that contains contiguous 16S-23S sequences. It was created from bacterial genomic data downloaded from RefSeq on May 21, 2019. A total of 5,551 “Reference” and “Representative” genomes, along with 13,634 other genomes assembled at the “Complete” and “Chromosome” levels were downloaded, for a total of 19,185 genomes. The NCBI Taxonomy database was used to annotate the RefSeq sequences. Target regions were extracted using a merger of NCBI’s GFF genome annotations and de novo annotations from Barrnap v0.8. Regions were defined by pairs of neighboring 16S and 23S genes that are between 2,000 and 8,500 base pairs. Since genomes frequently contain more than one 16S-23S gene pair, for example, Klebsiella genomes typically contains 8 separate 16S genes, the Athena database contains 55,964 individual 16S-23S regions. If a match for a read to a 16S-23S region in Athena, with errors included, is greater than 97%, and has a higher score than other matches, the read will be identified as the matching strain in the database. If a read matched equally well (or poorly) to multiple regions in the database, or if there is no match at the strain level, taxonomy will be reported at the highest level possible where an unambiguous call can be made. A similar cutoff is made at the species level, with a threshold of 95%. As a result, a novel *Klebsiella* read with a 96% identity to an existing strain would be reported as “*Klebsiella_oxytoca_unclassified*”, and a novel *Klebsiella* species will be reported as “*Klebsiella_unclassified*”.

### Athena Database Phylogenetic Categorization

A program was created to load the NCBI Taxonomy flat files into a searchable data structure. The data has a hierarchical structure, so a genome’s GFF3 file would typically provide a strain-level taxonomic classification for the genome, and the node representing that strain in the Taxonomy database would have a parent. The chain hierarchy can be followed to the tree root to determine the full taxonomic classification. To determine the taxonomic label for a genome in the Athena database, there were a few standardization issues to address. Most nodes in the Taxonomy database have multiple names, so the “best” name needed to be selected using heuristics. Also, some tree nodes have non-standard phylogenetic level categorizations such as “super-family” or “sub-genus”, so some harmonization was performed to map chains of nodes to the typical levels as much as possible: Kingdom, Phylum, Class, Order, Family, Genus, Species, Subspecies. This enabled the database output to assign taxonomic classification to specific levels such as “Level 6” while providing phylogenic consistency across samples. The program also modified the Taxonomy data to be more compatible with ‘mothur’(*24*); for example, spaces were replaced by underscores and parentheses were replaced with brackets. The program was used to create the “.taxonomy” file associated with the database that maps each sequence identifier to a multi-level phylogenetic classification.

### Mapping to Athena or SILVA Database

Database selection is integrated into the automated SBanalyzer pipeline. There are two choices in the drop-down menu for mapping reads, the Athena database and the SILVA database (*23*).

### Decontamination of Delftia reads

Almost all samples analyzed for StrainID amplicon sequences, including negative controls, appeared to have some level of *Delftia tsuruhatensis* contamination. *Delftia* is a known contaminant of laboratory water supplies (*26, 27*). *Delftia* could have been introduced in water used to dissolve the KOH for use in lysis solution, or in the TE used to elute DNA during sample preparation. The specificity of the StrainID amplicon enables the identification and removal of contaminating *Delftia* reads. *Delftia* reads called by SBanalyzer were confirmed by comparing *Delftia* sequences in the samples to all five 2,639 bp 16S-23S rRNA genes from *NZ_CP017420*.*1 Delftia tsuruhatensis strain CM13*, present in the Athena database, to determine how closely reads mapped. Reads were approximately 99.8% identical to the published genome, and similar across all contaminated samples, indicating that the *Delftia* contaminant was highly related to the sequenced strain. Interestingly, alignment of the reads in Jalview (*28*), showed that the variations from the reference genome mostly occurred between the 16S and 23S rRNA genes, between bases 1,700-2,050, where variation would not be expected to impact either the 16S or 23S rRNA genes, an indication that the contaminant is a novel *Delftia tsuruhatensis* strain. All reads from this previously unsequenced strain of *Delftia* were removed from the analysis.

### DADA2 Inference of Amplicon Sequence Variants

DADA2 is described for use with PacBio reads (*7*) and was installed as per instructions at: https://benjjneb.github.io/dada2/dada-installation.html.

FASTQ files were demultiplexed for each sample using SBanalyzer, with the “NoTrim” option in the SBanalyzer drop down menu. Reads assigned ‘Klebsiella’ taxonomy by SBanalyzer in the ‘.taxonomy’ file output were used as input for DADA2. The “R” script in Supplementary Data File S1 was used to perform read processing and specify the DADA2 parameters for ASV inference. The output includes two .png files, containing a sequence table heatmap and read length plots, a .FASTA file containing the Amplicon Sequence Variant sequences, and two R objects containing the workspace file and the dada2 output file.Demultiplexed FASTQ files were filtered on the basis of taxonomy assigned by SBanalyzer, selecting only reads at a desired taxonomic level (ex: “*Klebsiella*”) using a custom Python ‘readfinder’ script (Supplementary Data File S2).

The selected reads from each sample were primer trimmed, and filtered to reads within length range 1,900-3,000. The sequences within this length range were then dereplicated and passed to DADA2 to build an error model for read correction. The trimmed and filtered reads were analyzed manually via a histogram of the read lengths of all samples to identify peaks of read lengths that are likely to represent unique amplicons. The corresponding read length ranges (ex: the 2400-2405 bp range from each sample) were passed to dada2 and pooled for ASV inference. A sequence table of ASV abundance per sample was produced as part of the DADA2 output, and a heatmap was generated in R using the sequence table. Taxonomic information was added manually to the sequence table used to build the heatmap.

ASV sequences for *E. coli, Klebsiella* and *Enterobacter* are listed in Supplementary Data Files S5, S6, and S7, respectively

### rRNA phylogenetic analysis

*Klebsiella* StrainID sequences were exported from the Athena database and imported into Geneious Prime version 2020.1. Within Geneious the sequences were aligned using Clustal Omega (*29*) and manually curated. The phylogeny inferred using RAxML(*30*). The tree was annotated in iTOL (*31*).

### Determination of closely related type strains

Determination of closest type strain genomes was done in two complementary ways: First, all user genomes were compared against all type strain genomes available in the TYGS (*32*) database via the MASH algorithm, a fast approximation of intergenomic relatedness (*33*), and, the ten type strains with the smallest MASH distances chosen per user genome. Second, an additional set of ten closely related type strains was determined via the 16S rDNA gene sequences. These were extracted from the user genomes using RNAmmer (*34*) and each sequence was subsequently BLASTed (*25*) against the 16S rRNA gene sequence of each of the currently 11,300 type strains available in the TYGS database. This was used as a proxy to find the best 50 matching type strains (according to the bitscore) for each user genome and to subsequently calculate precise distances using the Genome BLAST Distance Phylogeny approach (GBDP) under the algorithm ‘coverage’ and distance formula d5 (*35*). These distances were finally used to determine the 10 closest type strain genomes for each of the user genomes.

### Pairwise comparison of genome sequences

All pairwise comparisons among the set of genomes were conducted using GBDP and accurate intergenomic distances inferred under the algorithm ‘trimming’ and distance formula d5. 100 distance replicates were calculated each. Digital DDH values and confidence intervals were calculated using the recommended settings of the GGDC 2.1(*35*).

### Phylogenetic inference

The resulting intergenomic distances were used to infer a balanced minimum evolution tree with branch support via FASTME 2.1.4 including SPR postprocessing (*36*). Branch support was inferred from 100 pseudobootstrap replicates each. The trees were rooted at the midpoint and visualized with PhyD3 (*37*).

### Type-based species and subspecies clustering

The type-based species were clustered using a 70% dDDH radius around each of the 32 type strains(*32*). Subspecies clustering was done using a 79% dDDH threshold (*38*).

## Data Availability

StrainID amplicon reads from each fecal sample and each Klebsiella isolate were uploaded to the NCBI BioProject database, with each sample as an indiv`idual file with the appropriate filename. All files were processed to remove contaminating Delftia reads. Annotated whole genome assemblies for K. oxytoca isolates were submitted to the NCBI BioProject database under BioProject accession number PRJNA608440.
StrainID amplicon data for this study is deposited under BioProjects PRJNA663638 and PRJNA663575. The accession numbers for the reads are SRR12647577-SRR12647615 and SRR12692773-SRR12692775.

## General

We are very grateful to Ben Callahan for reviewing the manuscript and for helpful comments and suggestions on DADA2 implementation for long read amplicons.

## Author contributions

JG and AM developed the hypotheses and designed the experiments with assistance from MC. MC performed the labwork. NL, EJ, SC, JG, AM, MD, DG analyzed the data. DF wrote the SBanalyzer software program. The manuscript was written by JG, AM and MD with substantial assistance from all the authors.

## Data and materials availability

If data are in an archive, include the accession number or a placeholder for it. Also include any materials that must be obtained through an MTA.

StrainID amplicon reads from each fecal sample and each *Klebsiella* isolate were uploaded to the NCBI BioProject database, with each sample as an indiv’idual file with the appropriate filename. All files were processed to remove contaminating *Delftia* reads. Annotated whole genome assemblies for *K. oxytoca* isolates were submitted to the NCBI BioProject database under BioProject accession number PRJNA608440.

StrainID amplicon data for this study is deposited under BioProjects PRJNA663638 and PRJNA663575. The accession numbers for the reads are SRR12647577-SRR12647615 and SRR12692773-SRR12692775.

## Supplementary Materials

**Supplementary Fig S1.**
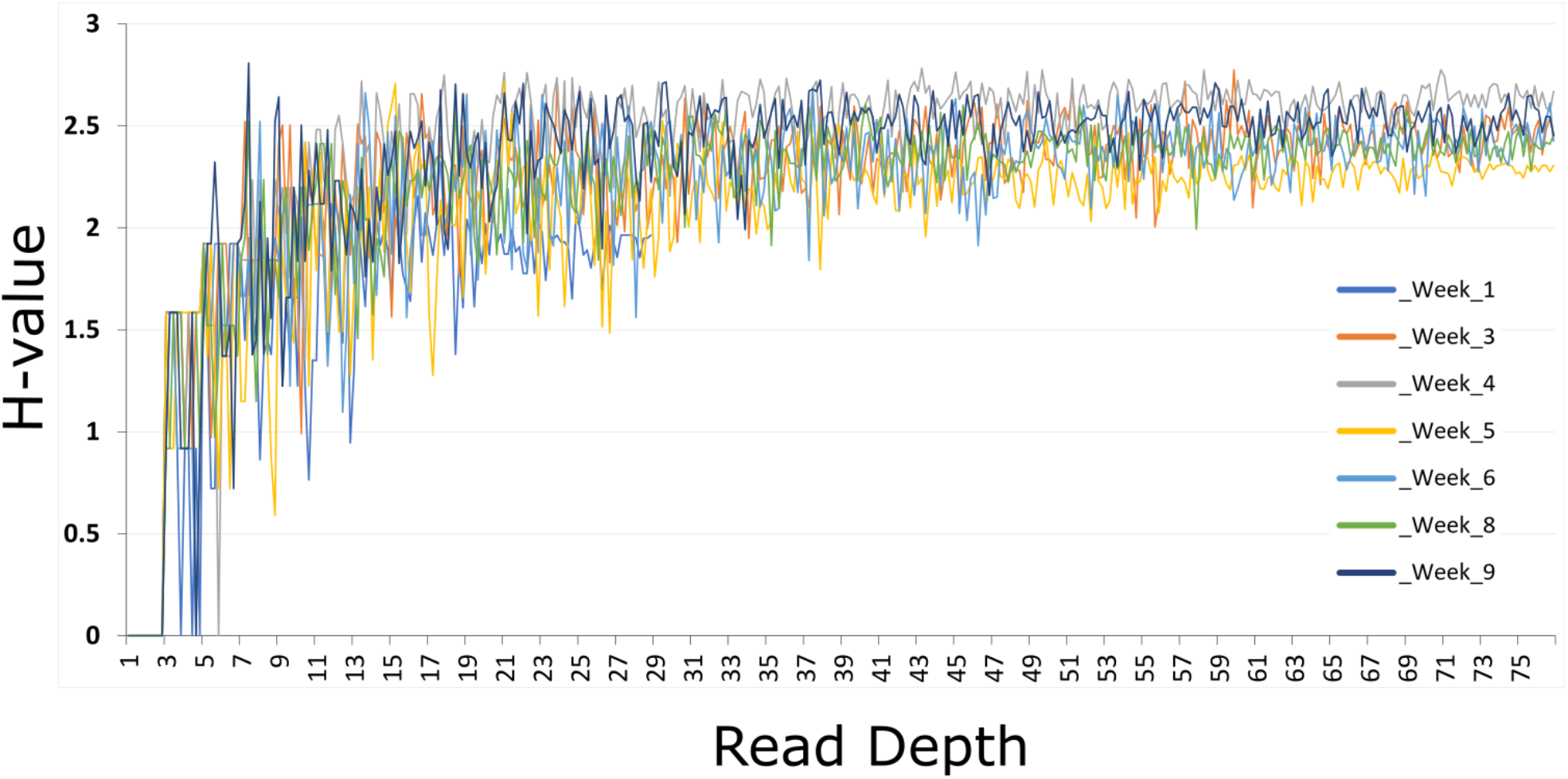
Longitudinal Shannon Diversity Plots for Twin Y. 10 iterations for H-value at read depth from 1-75 on the x-axis were calculated for each time point from Twin Y and plotted on the y-axis using QIIME2 (*40*)

**Supplementary Fig S2.**
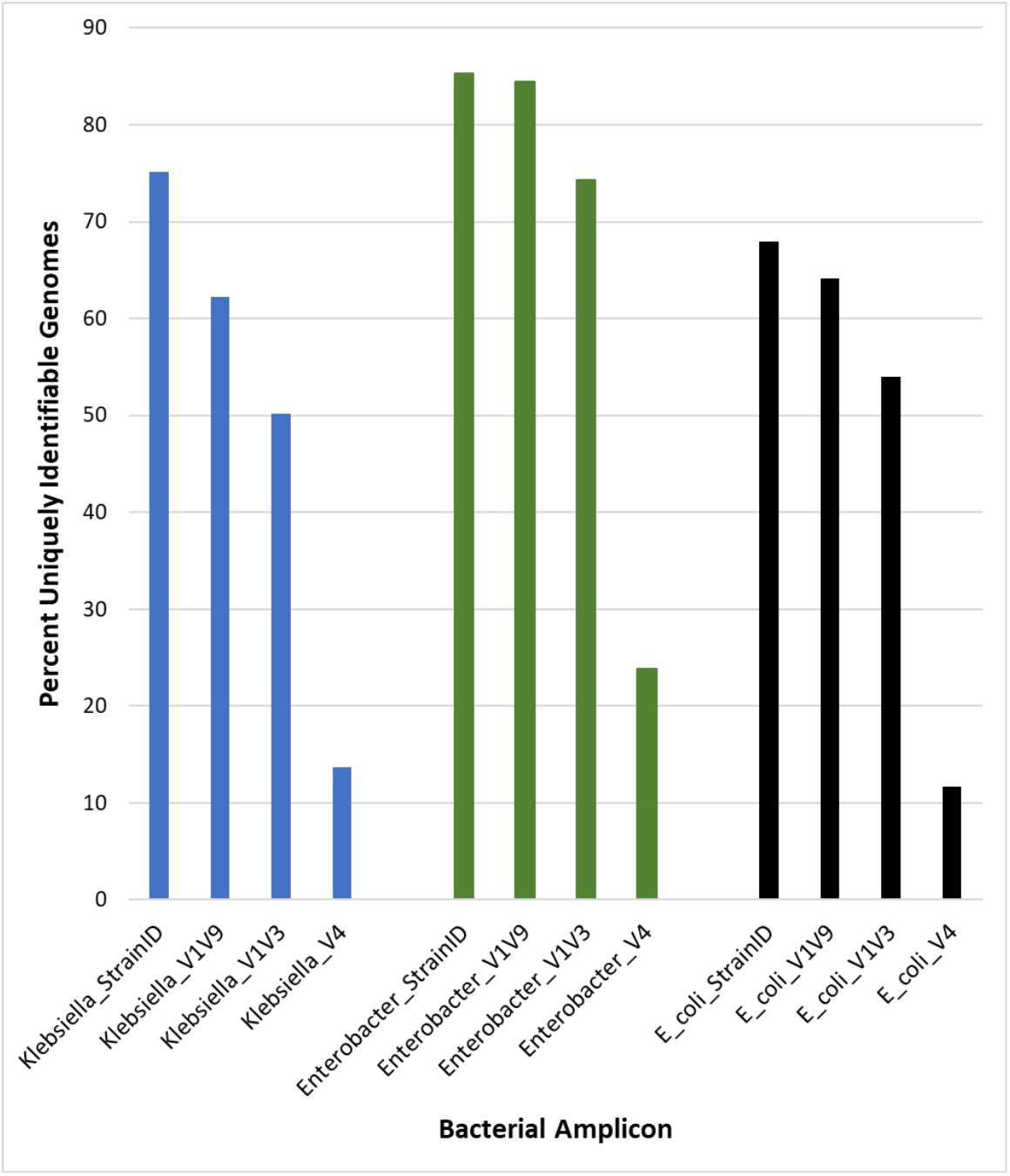
Uniquely Identifiable *Klebsiella, Enterobacter*, and *E. coli* Genomes in the Athena Database. Amplicon sequences were compared for 458 *Klebsiella*, 109 *Enterobacter*, and 187 *E. coli* genome entries in the Athena database. Each *Klebsiella* and *Enterobacter* genome ID contained up to eight contiguous 16S-23S gene pairs, *E. coli* contained up to seven, from which the sequences of the V4, V1-V3, V1-V9 and StrainID amplicons were extracted. A genome ID was considered unique if it contained either a unique amplicon, or a unique combination of amplicons. The percentage of uniquely identifiable genomes is shown on the y-axis, the taxonomy and amplicon type is shown on the x-axis.

**Supplementary Fig S3.**
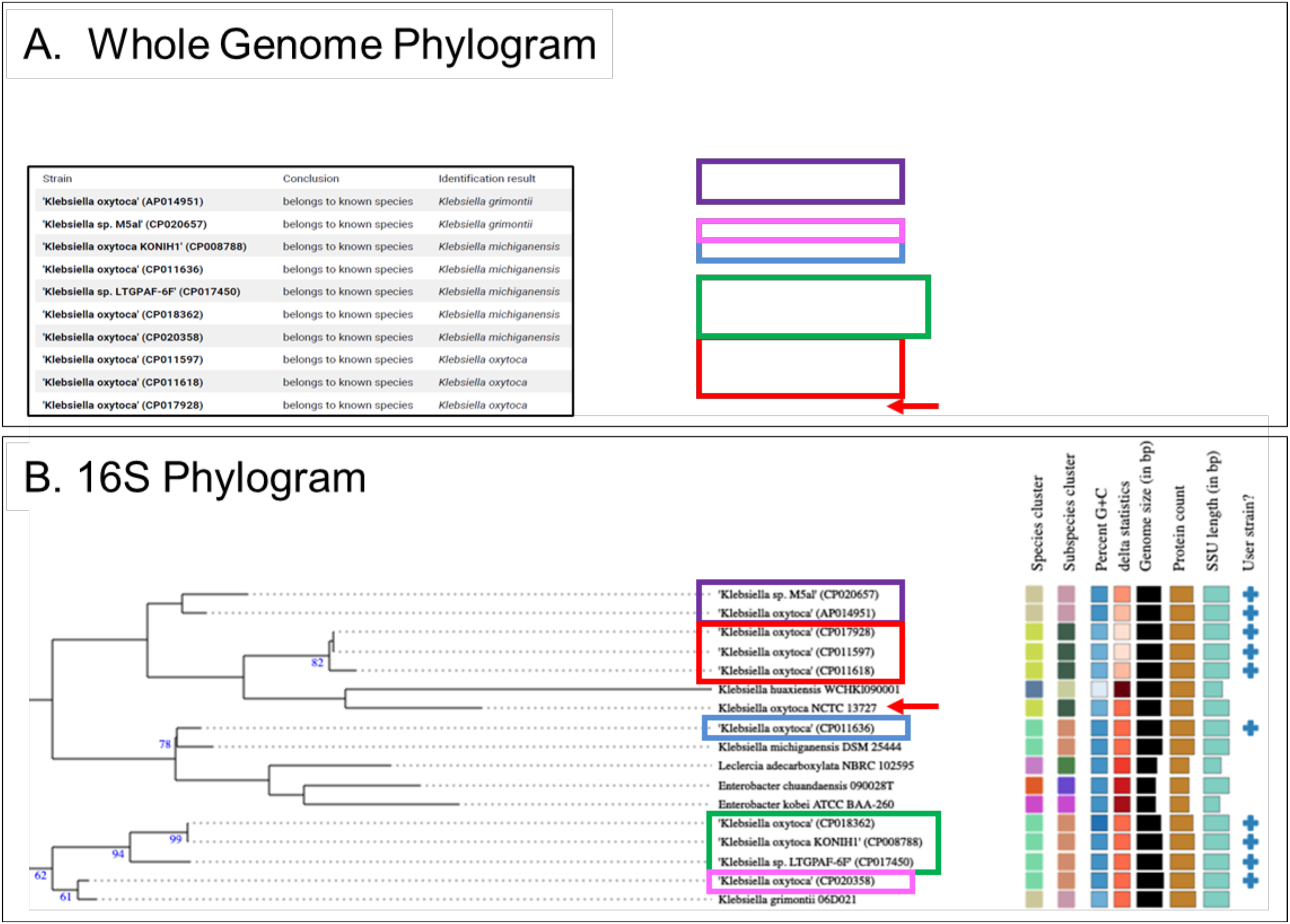
Whole Genome and 16S Phylogram Analysis. Ten *K. oxytoca* represented in the Athena database were compared with 11,300 strains in the TYGS database to identify closely related strains. The partial trees in the Figure include all branches needed to place the 10 ‘user strains’ selected from the Athena database. Colored boxes were used to indicate groups of genomes that sorted together. **(A)** Whole Genome Phylogram. Inset box indicates how the input Athena strains were identified by TYGS. The tree with detailed relationships was inferred with FastME 2.1.6.1 (*36*) from GBDP distances calculated from genome sequences. The branch lengths are scaled in terms of GBDP distance formula d5. The numbers above branches are GBDP pseudo-bootstrap support values > 60 % from 100 replications, with an average branch support of 85.5 %. The tree was rooted at the midpoint, not shown. **(B)** 16S Phylogram. Tree inferred with FastME 2.1.6.1 from GBDP distances calculated from 16S rDNA gene sequences. The branch lengths are scaled in terms of GBDP distance formula d5. The numbers above branches are GBDP pseudo-bootstrap support values > 60 % from 100 replications, with an average branch support of 53.3 %. The tree was rooted at the midpoint.

**Supplementary Fig S4.**
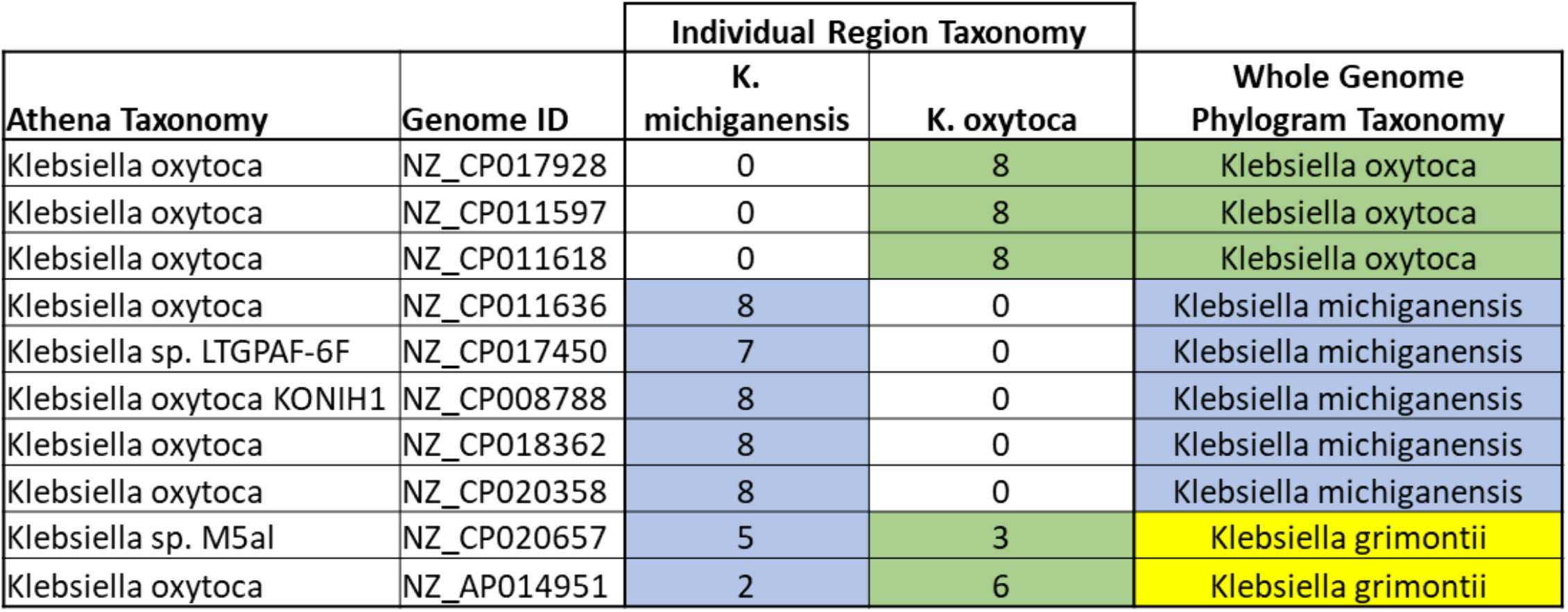
Individual 16S Region Taxonomies of Ten *Klebsiella* from Athena Database. Columns 1 and 2 show the Athena taxonomic assignment and the unique genome ID. Each of the 7-8 16S regions from each genome ID was mapped individually. 670 sequences were aligned using Clustal Omega (*29*) and manually curated before the phylogenic relationship was reconstructed using RAxML (*30*). The phylogenetic tree was annotated in iTOL (*31*), resulting in an individual region taxonomy for each that was either *K. michiganensis* or *K. oxytoca*, which were totaled in the appropriate columns. The Whole Genome Phylogram Taxonomy obtained from TYGS (*32*) is shown in the last column.

**Supplementary Data File S1**-DADA2 ‘R’ Script used to infer ASVs

**Supplementary Data File S2**-Python ‘readfinder.py’ script used to extract reads with taxonomies of interest from the data files

**Supplementary Data File S3**-SBanalyzer ‘.taxonomy’ file with read IDs and associated taxonomies assigned by SBanalyzer

**Supplementary Data File S4**-SBanalyzer ‘.groups’ file with read IDs and associated Sample Name Information

**Supplementary Data File S5** – FASTA file with *E. coli* Amplicon Sequence Variants

**Supplementary Data File S6** – FASTA file with *Klebsiella* Amplicon Sequence Variants

**Supplementary Data File S7** – FASTA file with *Enterobacter* Amplicon Sequence Variants

